# Microbiological Culture Versus 16S/18S Ribosomal RNA PCR-Sanger Sequencing for Infectious Keratitis: A Three-Arm, Diagnostic Cross-Sectional Study

**DOI:** 10.1101/2023.10.24.23297453

**Authors:** Yasmeen Hammoudeh, Lakshmi Suresh, Zun Zheng Ong, Michelle M. Lister, Imran Mohammed, D. John I. Thomas, Jennifer L. Cottell, Jennifer M. Holden, Dalia G. Said, Harminder S. Dua, Darren Shu Jeng Ting

## Abstract

**Purpose:** To compare the diagnostic performance of microbiological culture and 16S/18S polymerase chain reaction (PCR)-Sanger sequencing for infectious keratitis (IK) and to analyse the effect of clinical disease severity on test performance and inter-test concordance.

**Design:** A three-arm, diagnostic cross-sectional study.

**Subjects:** We included patients who presented with presumed bacterial/fungal keratitis to the Queen’s Medical Centre, Nottingham, UK, between June 2021 and September 2022.

**Methods/interventions:** All patients underwent simultaneous culture (either direct or indirect culture, or both) and 16S (pan-bacterial) / 18S (pan-fungal) ribosomal RNA (rRNA) PCR-Sanger sequencing. The bacterial/fungal genus and species identified on culture were confirmed using matrix-assisted laser desorption/ionization-time-of-flight mass spectrometry. Relevant clinical data were also collected to analyze for any potential clinico-microbiological correlation.

**Main outcome measures:** Diagnostic yield, test accuracy (including sensitivity and specificity), and inter-test agreement [including percent agreement and Cohen’s kappa (*k*)].

**Results:** A total of 81 patients (86 episodes of IK) were included in this study. All organisms identified were of bacterial origin. Diagnostic yields were similar among direct culture (52.3%), indirect culture (50.8%), and PCR (43.1%; p=0.13). The addition of PCR enabled a positive diagnostic yield in 3 (9.7%) direct culture-negative cases. Based on composite reference standard, direct culture had the highest sensitivity (87.5%; 95% CI, 72.4-95.3%), followed by indirect culture (85.4%; 95% CI, 71.6-93.5%) and PCR (73.5%; 95% CI, 59.0- 84.6%), with 100% specificity noted in all tests. Pairwise comparisons showed substantial agreement among the three tests (percent agreement=81.8-86.2%, Cohen’s *k*=0.67-0.72). Clinico-microbiological correlation demonstrated higher culture-PCR concordance in cases with greater infection severity.

**Conclusions:** This study highlights a similar diagnostic performance of direct culture, indirect culture and 16S rRNA PCR for bacterial keratitis, with substantial inter-test concordance. PCR serves as a useful diagnostic adjuvant to culture, particularly in culture-negative cases or those with lesser disease severity (where culture-PCR concordance is lower).

## INTRODUCTION

Infectious keratitis (IK), or commonly known as corneal infection, is the primary cause of corneal blindness globally.^1^ Depending on the geographical and temporal variations, the incidence of IK has been estimated to be 2.5-799 cases per 100,000 people annually, with a significantly higher incidence and prevalence observed in low- and middle-income countries.^1,2^ Timely and accurate diagnosis followed by appropriate treatment is key to achieving a good clinical outcome.^3^ Studies have shown that visual outcome, corneal healing time and treatment success of IK are significantly influenced by the initial presenting severity of the infection.^4–6^

IK is primarily diagnosed on clinical grounds, supplemented by microbiological investigations and/or imaging test.^3^ Corneal sampling for microscopy (with staining), microbiological culture and susceptibility testing are the current mainstay for IK diagnosis as it can determine the causative organisms and their antimicrobial susceptibility and/or resistance. Microbiological culture can be performed using either direct culture or indirect culture methods, with both showing similar diagnostic yields in many studies.^7–10^ The advantage of indirect culture is that it is clinically less arduous since it only requires one sampling by the front-line ophthalmologists/doctors (instead of multiple samplings as required in direct culture). The sample is then inoculated in a transport medium for further sub-inoculation on multiple agar plates at a microbiological laboratory, making it a more efficient and practical test in a hectic clinical environment.

However, microbiological culture (in the setting of IK) is hindered by its variably low diagnostic yield due to considerably lower infectious biomass (when compared to systemic infections),^11^ potential inadequate sampling, prior use of antimicrobials,^12^ and poor growth of unusual pathogens (which can be further affected by a wrong choice of culture medium).^3^ In addition, a positive culture result may take 2-3 days (and up to 14 days for some slow-growing pathogens) to become available, negatively impacting on the management of IK. As a result, newer molecular techniques such as polymerase chain reaction (PCR) have been increasingly explored and used for diagnosing IK.^3,13,14^

PCR is a highly sensitive and time-efficient diagnostic test which can typically generate a positive (or negative) result within a few hours, making it an ideal diagnostic modality for IK. Among various assays and techniques, broad-range 16S/18S ribosomal RNA (rRNA)-based PCR has emerged as one of the most popular methods.^3,14–16^16S rRNA is a major component of the 30S small subunit present in all prokaryotes, including bacteria. It contains multiple highly conserved regions (useful for universal PCR priming) and nine variable regions (V1-V9; pertinent for phylogenetic analysis and taxonomic resolution).^17^ On the other hand, 18S rRNA (a core component of 40S small subunit) is highly conserved by eukaryotes, including fungi and Acanthamoeba, and it similarly contains nine variable regions (V1-V9).^18^ Therefore, targeting the highly conserved, universal 16S/18S rRNA gene should theoretically identify almost any bacterial or fungal pathogens, including those that are difficult to grow in conventional culture media. In addition, PCR is able to detect microbial DNA in viable and non-viable organisms and is not affected by prior use of antimicrobials.^19^

In our recent 12-year Nottingham Infectious Keratitis Study,^20^ we observed a low culture yield (37.7%) in IK. As a result, 16S/18S rRNA PCR was introduced to the local clinical care pathway in June 2021 as part of the standard IK workup to improve the diagnostic yield. To date, there were only two studies conducted in the UK specifically compared the diagnostic performance between culture and PCR for IK.^14,21^ In view of the apparent gap in the literature, this study aimed to compare the performance among direct culture, indirect culture and 16S/18S rRNA PCR for IK and to examine the concordance among these tests.

## METHODS

### Study Design

This was a monocentric, three-arm, diagnostic cross-sectional study. The study was approved by the clinical governance team at the Nottingham University Hospitals NHS Trust as a clinical quality improvement project (Ref: 21-135C) with an aim to improve culture yield for IK.^20^ The study was conducted in accordance with the tenets of the Declaration of Helsinki. As corneal sampling formed part of the standard clinical care for IK, no additional informed patient consent (over and above the usual verbal consent for corneal sampling) was required. The study was reported according to the Standards for Reporting of Diagnostic Accuracy Studies (STARD) guideline for completeness and transparency.^22^

### Case Identification

Consecutive patients who presented with clinically suspected BK or FK and underwent corneal sampling for conventional microbiological culture (direct culture, indirect culture or both) and pan-bacterial (16S rRNA) / pan-fungal (18S rRNA) PCR at the Queen’s Medical Centre (QMC), Nottingham, UK, between June 2021 and September 2022 were prospectively included. The diagnosis of IK was made by at least one corneal consultant and/or fellow based on clinical features, microbiological results, and/or corneal imaging. In patients where repeated samplings were performed at different time points, each episode was included and analyzed separately as the primary aim of this study was to compare the diagnostic accuracy and agreement among the tests. Acanthamoeba and viral keratitis cases were excluded from this study.

### Data Collection

All microbiological results, including culture and PCR, were collected and stored within the Nottingham local microbiological database.^20,23^ Other relevant clinical data, including demographic factors, risk factors, clinical characteristics, corrected-distance-visual-acuity (CDVA), management, outcomes, and complications, were collected from the local electronic health records and were collated using a standardized excel proforma for secondary analysis for any potential clinico-microbiological correlation. For patients that underwent repeated sampling, demographic and clinical data were only obtained from the first IK episode of each patient to avoid duplication, but all IK episodes were analyzed from the standpoint of microbiological tests as mentioned above. For bilateral IK cases, only one eye (i.e. the more severely affected eye) was included in this study. Similar to our previous studies,^4,5^ the size of ulcer (including epithelial defect and infiltrate) was categorized as small (<3mm), moderate (3-6 mm), and large (>6mm), based on maximum linear dimension. The location of the ulcer was classified into peripheral (the entire ulcer was within 3 mm from the limbus), paracentral (between peripheral and central location), and central (any part of the ulcer affecting the visual axis).

### Corneal Sampling Procedure for Culture and PCR

As per the local departmental protocol, corneal sampling was performed when one or more of the following criteria were met: infiltrate size >1mm, centrally located ulcer, significant anterior chamber activity / hypopyon, bilateral cases, atypical presentation, or unresponsive to antimicrobial treatment.^4^ In refractory IK cases, all antimicrobial treatment were withheld for >24 hours before corneal re-sampling was performed. In view of the low culture yield (37.7%) demonstrated in our previous study,^20^ corneal sampling with flocked swabs [FLOQSwab (product code: 537CS01); Copan Italia s.p.a., Brescia, Italy) was introduced to replace needles/blades to improve culture and PCR yield. These flocked swabs have a perpendicularly sprayed-on nylon fiber coating at the tip, which could increase microbial uptake and release (hence a better yield).^9,24^

As previous studies had shown that the order of corneal sampling did not influence the diagnostic yield,^9,14^ a standardized approach and order of corneal sampling was adopted in this study (**Figure 1**). The first corneal sampling was performed using a nylon flocked swab with subsequent inoculation in a tube containing 1 ml of modified Amies transport medium [eSwab kit (product code: SS451), Sterilin, Appleton Woods Ltd, Birmingham]. This was then utilized for downstream processing for indirect culture (within 24 hours) and 16S/18S PCR analysis (within 24-48 hours). Four subsequent corneal samplings were performed using four single-use corneal FLOQSwabs and inoculated onto the agar plates (i.e., the direct culture method), including one chocolate agar plate, one blood agar plate, one fastidious anaerobe agar plate, and one Sabouraud dextrose agar plate.^4^

**Figure 1.**
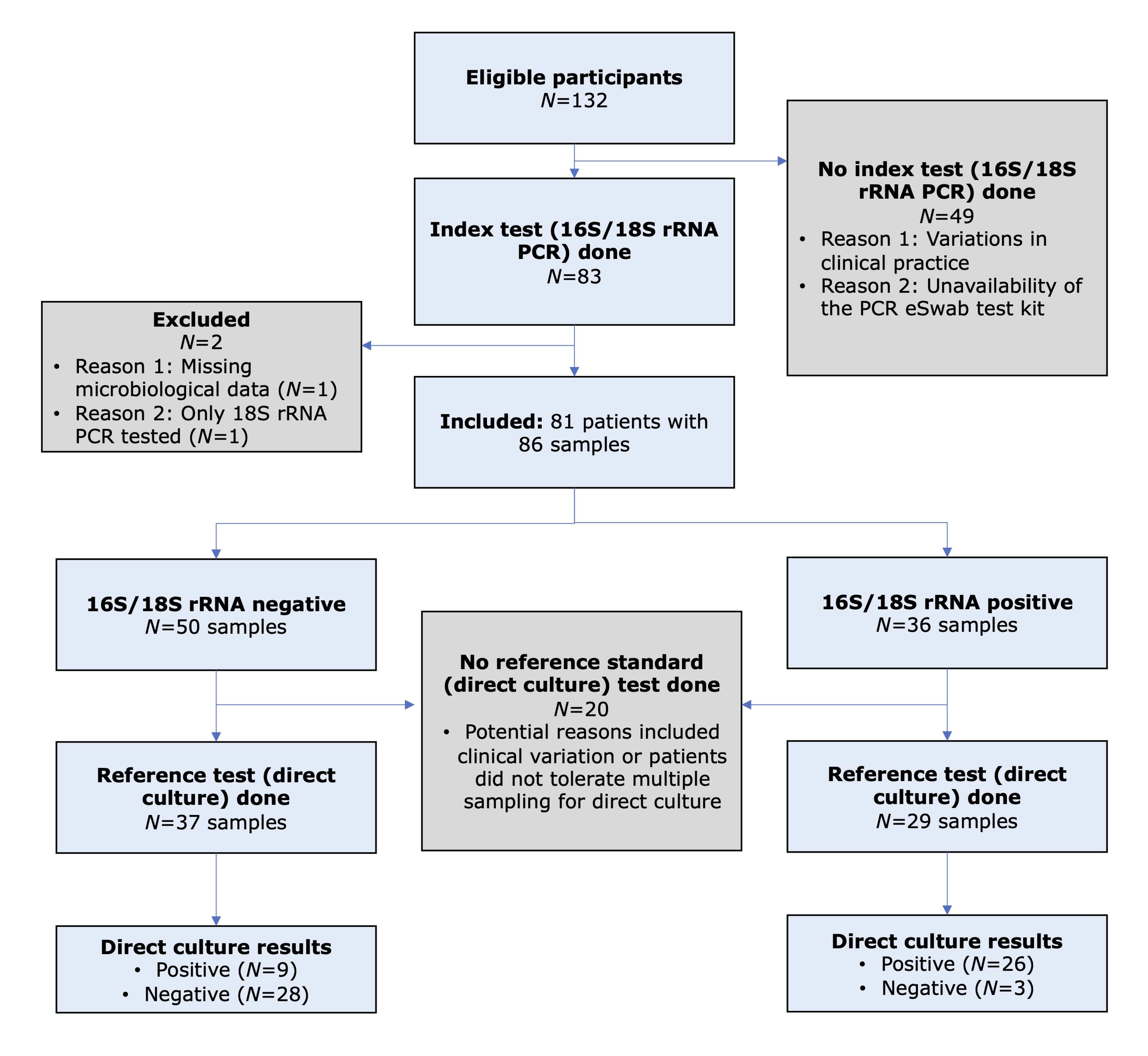
Flow diagram of the study, based on the Standard for Reporting Diagnostic Accuracy (STARD) guideline, comparing the diagnostic performance of 16S/18S ribosomal RNA polymerase chain reaction (PCR; index test) and direct culture (reference standard) for diagnosing presumed bacterial or fungal keratitis.

All corneal samples were sent to the in-house microbiological laboratory at the QMC, Nottingham, UK, for further processing. For indirect culture, 80 µl of the inoculated Amies liquid medium were obtained after vortexing the swab and liquid medium, aliquoted equally and inoculated onto four agar plates (same as the direct culture method; 20 µl per plate). All culture plates (both direct and indirect culture) were incubated for at least five days (and up to three weeks for suspected fungal keratitis) within the microbiological laboratory. Bacterial identification in cultures were confirmed by matrix-assisted laser desorption/ionization-time-of-flight mass spectrometry [(MALDI-TOF MS); Bruker, Coventry, UK] with a score of >2.0, indicating secure genus and probable species identification. For pan-bacterial (16S) / pan-fungal 18S rRNA PCR analysis, 500 µl was obtained from the inoculated Amies medium. Of this, a 200 µl aliquot was taken and added to lysozyme (25 µl at 100 mg/ml) and lysostaphin (10 µl at 1 mg/ml) and incubated for 30 minutes at 37 °C. Afterwards an aliquot of 40 µl of Proteinase K is added along with 200 µl of lysis buffer instructions and subjected to heat-treatment according to the manufacturer’s instructions (Qiagen). Nucleic acid is then extracted using the QiaSymphony Bio-Robot (Qiagen) using the QiaSymphony DSP DNA Mini Kit according to manufacturer’s instructions with a final DNA eluate of 200 µl.

### Pan-Bacterial 16S rRNA Gene Assay

A single-round amplification was performed using appropriate primers as described by Relman et al.^25^ Each reaction mix consisted of 14 µl of 2 x MyTaq Mix (Meridian scientific), 0.5 µl of Evagreen Sybr Green (BIOTIUM), 5 µl of Nuclease free water (Severn Biotech), and 1 µM of the forward and reverse primers (16S Uni P-F: AGAGTTTGATCATGGCTCAG; 16S Uni P-R:ACCGCGGCTGCTGGCAC), and a 5 µl aliquot of extracted DNA was added to this mix. The reaction mixes were subjected to thermal cycling on a magnetic induction real-time cycler with a programme of initial heating for 95 °C for 1 min 45s, followed by 30 cycles of 95 °C for 20 s, 55 °C for 20s and 72 °C for 20s. Melt profiles were scored against positive controls and any potential positive PCR reactions were electrophoresed through a 3 % agarose gel containing ethidium bromide and bands were visualized by UV transillumination before being sequenced. Similar to previous studies,^26^ the total PCR cycle number was set at 30 (but not higher) to reduce false positive result and improve positive predictive value of this broad-range assay.

### Pan-Fungal 18S rRNA Gene Assay

A single-round amplification was performed using appropriate universal primers that targeted the fungal 18S internal transcribed space 1 (ITS1) region of the rRNA gene cluster (as described by Lau et al.).^27^ Each reaction mix consisted of 25 µl of 2 x MyTaq Mix (Meridian scientific), 5 µl of nuclease free water (Severn Biotech), and 1 µM of the forward and reverse primers (Fun1: TCCGTAGGTGAACCTGCGG; Fun2: GCTGCGTTCTTCATCGATGC), and a 20 µl aliquot of extracted DNA was added to this mix. The reaction mixes were subjected to thermal cycling with a programme of initial heating for 95 °C for 1 min 45 s, followed by 40 cycles of 95 °C for 20 s, 60 °C for 20 s and 72 °C for 20 s. PCR reactions were electrophoresed through a 3 % agarose gel containing ethidium bromide, and bands were visualized by UV transillumination.

### Sanger Sequencing and Phylogenetic Typing

Amplicons were purified using Sureclean Plus (Bioline) according to manufacturers instructions and eluted in nuclease free water. BigDye Terminator v3.1 Cycle Sequencing Kit (Thermo Fisher Scientific) was used to set up Sanger sequencing reactions according to the following mix: 0.5 µl of BigDye Terminator v3.1 Ready Reaction Mix, 1 µl of 5x Sequencing Buffer, 1 µl of 5 µM primer, 1 µl purified DNA template, and 6.5 µl nuclease free water. The reaction mixes were subjected to thermal cycling with a programme of initial heating for 96 °C for 1 min, followed by 25 cycles of 96 °C for 10 s, 50 °C for 5 s and 60 °C for 4 mins. BigDye Sequencing reactions were then cleaned up using BD Xterminator solution (Thermo Fisher Scientific) according to manufacturer’s instructions. Sanger sequencing was performed using the 3500XL Genetic Analysers (Applied Biosystems) using a 50 cm array. Sequence data was basecalled and trimmed using SeqA software (Applied Biosystems) and aligned using ChromasPro sequencing software (Technelsium). Unambiguous taxonomic assignment of sequence data was performed using Basic Local Alignment Search Tool (BLAST, NCBI) against the nr/nt database.

### Power and Sample Size Calculation

The sample size of this 3-arm comparative study was calculated based on an equivalence study design, assuming a 40% diagnostic yield for each test, with a minimum detectable effect set at 20% and an equivalence margin set at 20% (power=80% and p<0.05). Based on this calculation, a total of 61 samples per each test group were required.

### Statistical Analysis

Statistical analysis was conducted using SPSS v28.0 (IBM SPSS Statistics for Windows, Armonk, NY, USA). For descriptive and analytic purposes, patients were divided into microbiological-positive (i.e. patients with a positive result on any of the three tests) and microbiological-negative (i.e. patients with negative results on all three tests) groups and were analyzed for any potential clinico-microbiological correlation. Comparison between groups was conducted using Pearson’s Chi square or Fisher’s Exact test for categorical variables, and T test or Mann-Whitney U test for continuous variables where appropriate. Direct culture was the reference test whereas 16S/18S rRNA PCR and indirect culture were the primary and secondary index tests, respectively. All continuous data were presented as mean ± standard deviation (SD) and/or 95% confidence interval (CI).

The main outcome measures were the diagnostic performance (including yield, sensitivity, and specificity) and the inter-test agreement. Sensitivity and specificity of the tests were estimated using two reference standards, including direct culture alone and composite reference standard (CRS), defined as at least one positive result for any test (**Supplementary Table 1**).^21^ Vassar Stats (vassarstats.net) was used to calculate the 95% confidence intervals (CIs) for the sensitivity and specificity of each test. Cochran’s Q test was used to compare the diagnostic performance among the three tests at the positive microbiological detection level (i.e. based on positive or negative microbiological results). Percent agreement and Cohen’s kappa (κ) were used to examined inter-test pairwise agreement between any two tests at the organism genus level, and κ was interpreted as: (1) poor: 0.00-0.20; (2) fair: 0.21-0.40; (c) moderate: 0.41-0.60; (d) substantial: 0.61-0.80; and (e) almost perfect: 0.81-1.00.^28,29^ In addition, potential influencing clinical factors on inter-test agreement (for microbiological-positive cases) between direct culture and PCR was also analyzed. A p-value of <0.05 was considered statistically significant.

## RESULTS

### Patient and Clinical Characteristics

A total of 81 patients (with 86 IK episodes) were included (**Figure 2**). The mean patient age was 49.8 ± 22.7 years, and 41 (50.6%) patients were female. Of the 81 patients, four underwent repeated sampling, one of which had two repeated samplings. Overall, 47 (58.0%) and 34 (42.0%) patients were microbiological-positive and microbiological-negative, respectively. Similar demographic factors and clinical characteristics were observed in both microbiological-positive and microbiological-negative groups [all p-values >0.05, except for the presence of hypopyon (p=0.036); **Table 1**]. Contact lens wear (43, 53.1%), ocular surface disease (34, 42.0%), and use of topical steroids (14, 17.3%) were the three most common risk factors. The mean presenting CDVA was 0.93 ± 0.93 logMAR. The majority of the ulcers were of small epithelial defect size (48, 59.3%), small infiltrate size (51, 63.0%), paracentrally located (37, 45.7%), and absence of hypopyon (65, 80.2%).

**Figure 2.**
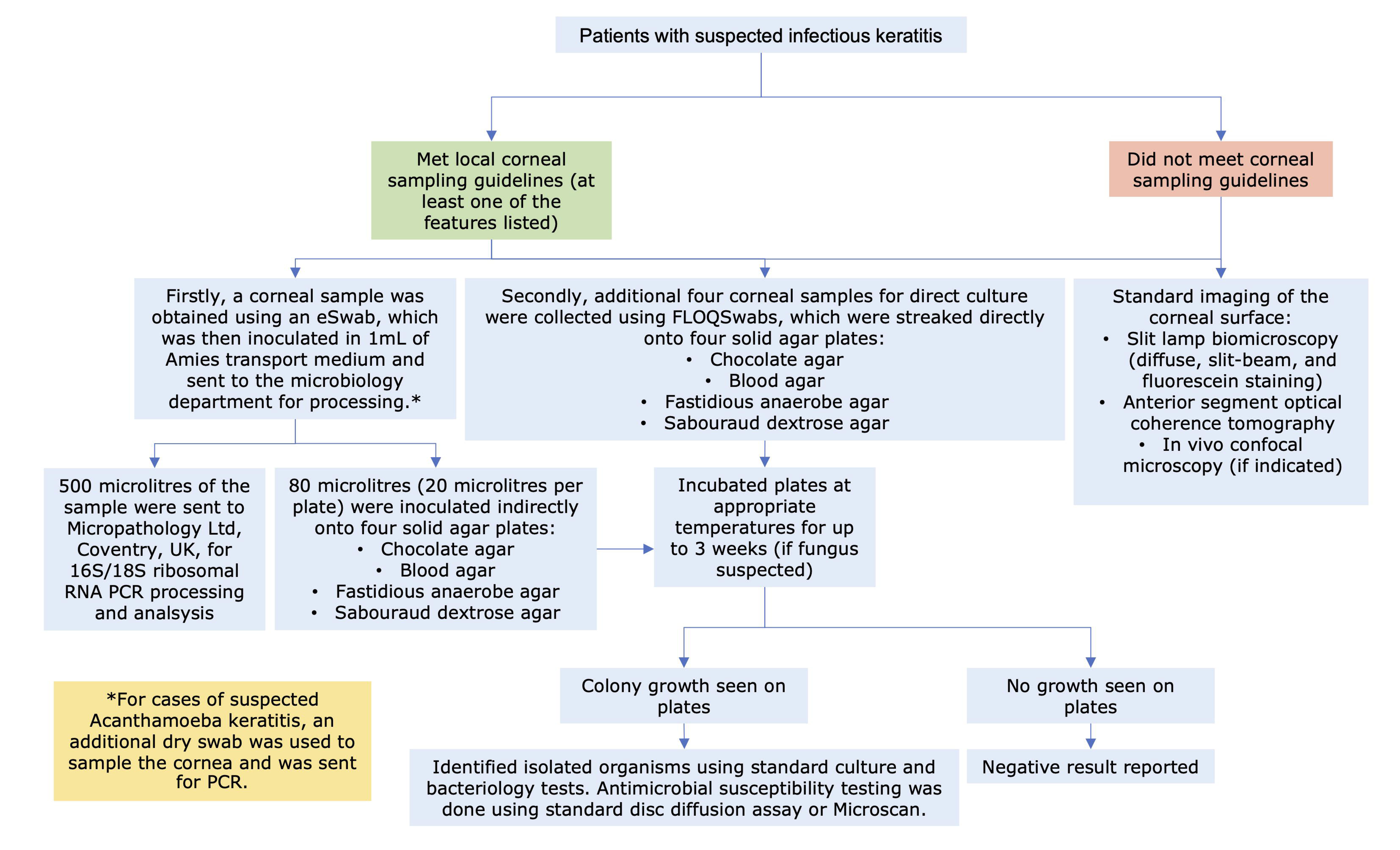
Flow diagram detailing the study methodology on the diagnostic pathway of infectious keratitis at the Queen’s Medical Centre, Nottingham, UK.

### Diagnostic Yield and Causative Organisms

Of the 86 included cases, 66 (76.7%), 85 (98.8%), and 86 (100.0%) cases underwent direct culture, indirect culture, and PCR, with 65 (75.6%) cases having had all three tests performed. The diagnostic yield for direct culture, indirect culture and PCR was 53.0%, 48.2%, and 41.9%, respectively. When considering only cases that were investigated by all three tests (n=65), the diagnostic yield changed slightly to 52.3% (34/65), 50.8% (33/65), and 43.1% (28/65) for direct culture, indirect culture, and PCR (**Figure 3**), with no significant difference among the tests (Cochran’s Q test, p=0.13). When using direct culture as the reference standard, the addition of PCR was able to improve the diagnostic yield of direct culture-negative cases by 9.7% (3 of the 31 cases).

**Figure 3.**
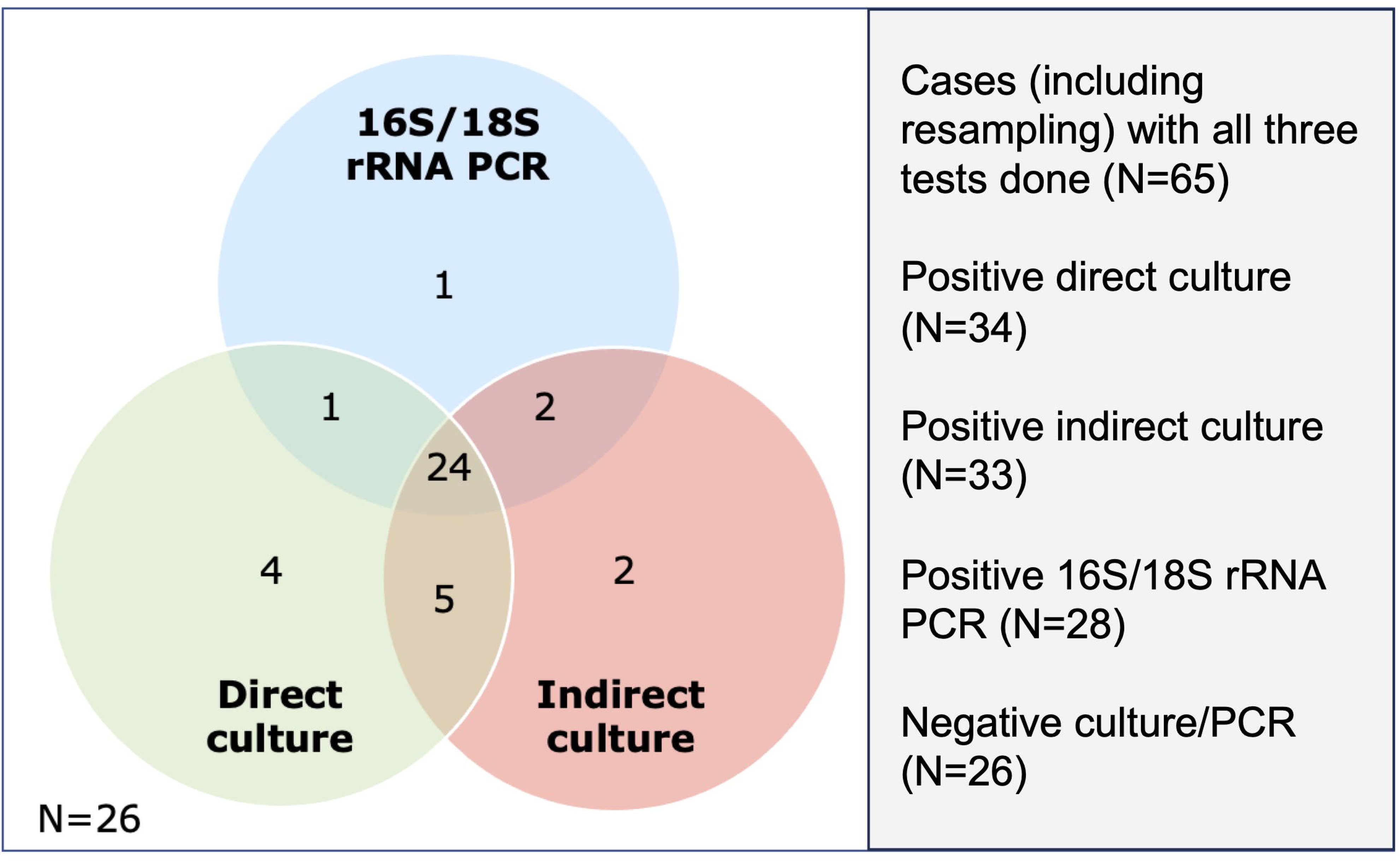
Venn diagram demonstrating the number of positive cases detected on three microbiological tests, including direct culture, indirect culture, and 16S/18S rRNA polymerase (PCR).

A total of 66 causative organisms (all bacteria and no fungus) were identified (**Table 2**). Coagulase-negative staphylococci (15, 22.7%), *Pseudomonas aeruginosa* (14, 21.2%), and *Propionibacterium spp.* (13, 19.7%) were the most common organisms isolated. PCR detected 36 organisms, including three organisms that were not identified on either direct or indirect culture, namely *Acinetobacter spp., Bacillus spp.*, and *Staphylococcus capitis* (a type of CoNS). There was a significantly lower proportion of *Propionibacterium spp.* identified in the PCR group (2.8% of all organisms) when compared to the direct culture (12.2%) and indirect culture (22.2%) groups (p=0.030). There were 14 (17.3%) cases of polymicrobial infection when the results of all three tests were considered together.

Seven (8.1%) cases were associated with the use of topical antimicrobial treatment (mainly chloramphenicol) before corneal sampling. A causative organism was identified in two (28.6%) of the cases, including a case of *P. aeruginosa* (positive on all three methods) and a case of *Propionibacterium spp.* (only positive on indirect culture). The remaining five cases had both negative culture and PCR results, suggesting similar diagnostic performance between culture and PCR in cases with prior use of antimicrobial.

### Diagnostic Performance

#### Direct culture reference standard

When using direct culture as the reference standard, the sensitivity and specificity of indirect cultures (n=65) were 85.3% (95% CI 68.2-94.5%) and 87.1% (95% CI 69.2-95.8%), respectively (**Table 3** and **Figure 4**). On the other hand, the sensitivity and specificity of 16S/18S rRNA PCR (n=66) were 74.3% (95% CI 56.4-86.9%) and 90.3% (95% CI 73.1- 97.5%), respectively (**Table 3** and **Figure 4**).

**Figure 4.**
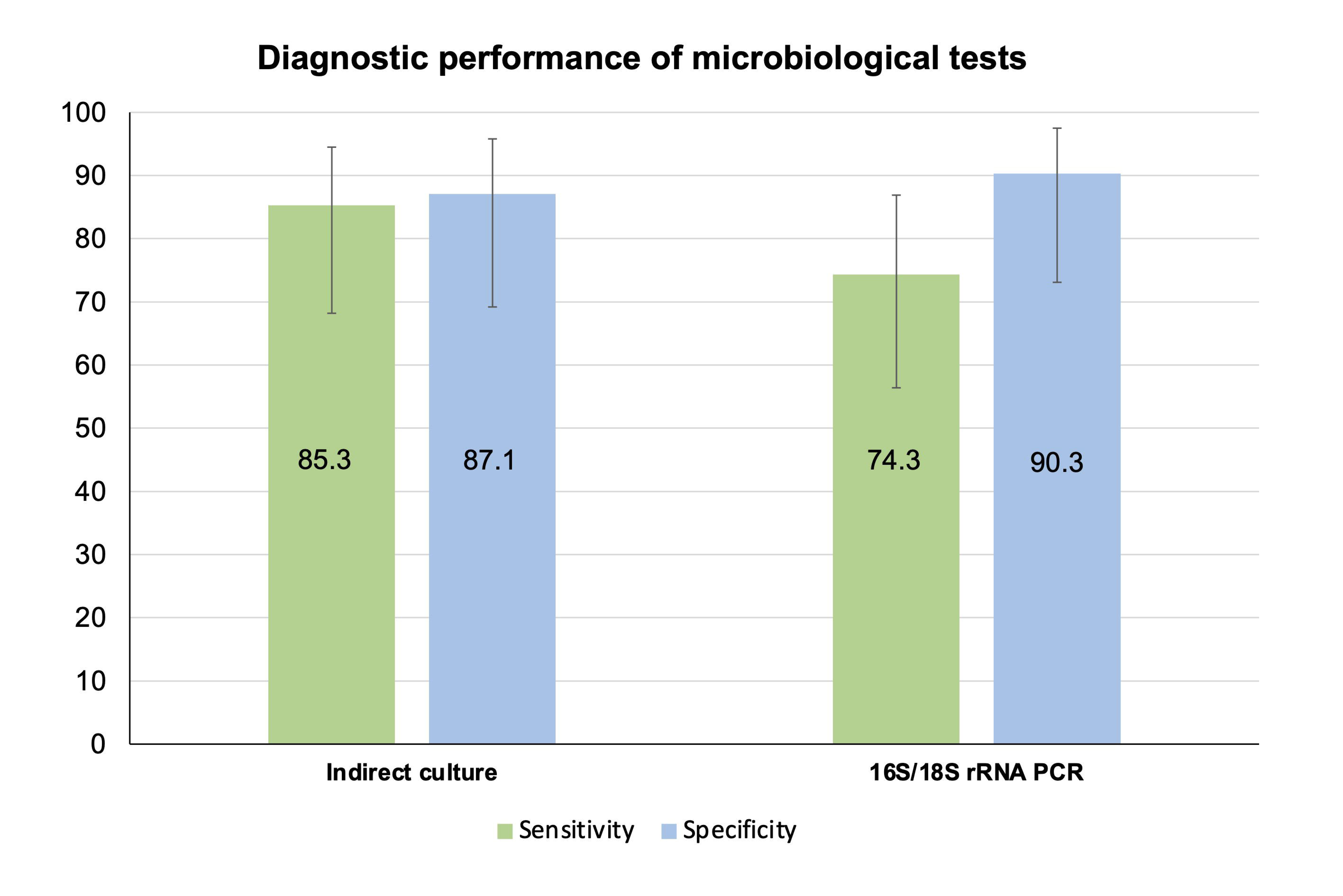
Diagnostic performance of indirect culture and 16S/18S rRNA PCR-Sanger sequencing, using direct culture as the reference standard.

#### Composite reference standard (CRS)

Based on the CRS, the sensitivity of direct culture (n=66), indirect culture (n=85), and PCR (n=86) was 87.5% (95% CI 72.4-95.3%), 85.4% (95% CI 71.6-93.5%), and 73.5% (95% CI 59.0-84.6%), respectively (**Table 4** and **Figure 5**). The specificity of all three tests was 100% (95% CI 84.0-100.0% for direct culture and 95% CI 88.3-100.0% for indirect culture and PCR).

**Figure 5.**
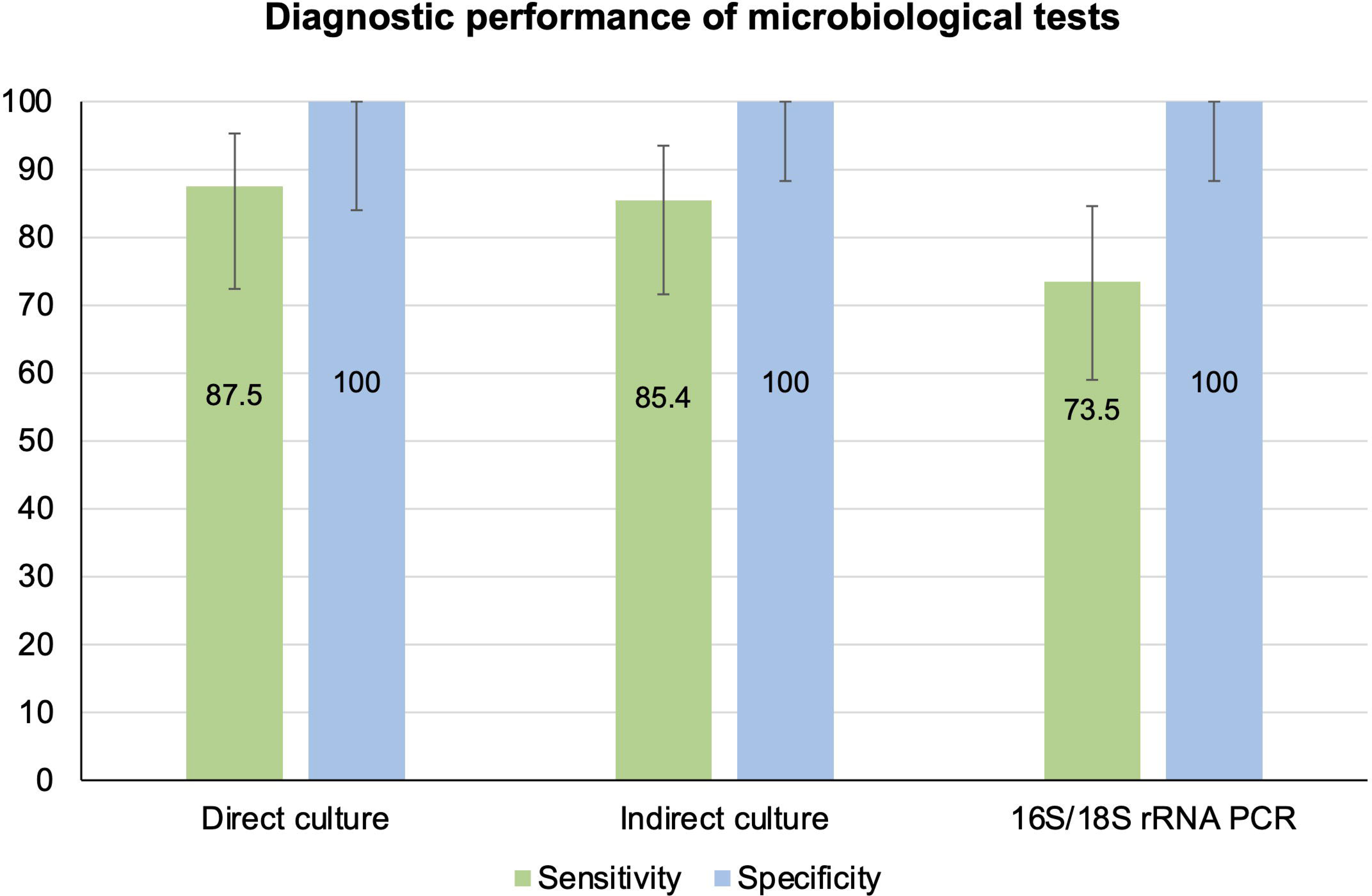
Diagnostic performance of direct culture, indirect culture, and 16S/18S rRNA PCR-Sanger-sequencing, using composite reference standard as the reference standard.

### Agreement between Microbiological Investigations

#### At microbiological detection level

Pairwise comparison between any two investigations at the microbiological detection level showed the greatest agreement/concordance with direct culture-indirect culture (86.2%), followed by indirect culture-PCR (85.9%) and direct culture-PCR (81.8%). Cohen’s kappa analysis showed a substantial pairwise agreement with any of the two microbiological tests (mean k=0.67-0.72; **Table 5**).

#### At organism’s genus level

When considering at the organism’s genus level, the concordance between any two tests remained similarly good. The concordance was the greatest with direct culture-indirect culture (84.6%), followed by indirect culture-PCR (82.4%) and direct culture-PCR (77.3%). There was a substantial agreement between direct culture-indirect culture (*k*=0.69 ± 0.09; 95% CI: 0.52-0.87), and indirect culture-PCR (*k*=0.62 ± 0.08; 95% CI: 0.46-0.79).

Comparison between direct culture and PCR showed moderate agreement (*k*=0.55 ± 0.10; 95% CI: 0.36-0.74; **Table 5**).

#### Potential influencing factors for inter-test agreement

Based on direct culture and PCR results in microbiological positive (either culture- or PCR-positive) IK cases, we found that culture-PCR concordance was more likely to be achieved in cases with more severe infection (**Table 6**). Culture-PCR matched cases had a worse mean presenting CDVA (1.29 logMAR vs. 0.62 logMAR; p=0.040), larger epithelial defect (>3mm; 74.0% vs. 36.4%; p=0.16), larger infiltrate (>3mm; 40.0% vs. 25.0%; p=0.48), central location (48.0% vs. 27.3%; p=0.30), presence of hypopyon (42.3% vs. 16.7%; p=0.16) than culture-PCR unmatched cases, though statistical significance was not achieved in most parameters likely due to a type 2 error (i.e. small sample size).

## DISCUSSION

Timely diagnosis and treatment serve as the key to achieving good outcomes in IK. To overcome the inherent limitations of microbiological culture, PCR test has increasingly gained traction for diagnosing IK in recent years in view of their high sensitivity and high specificity.^3^ So far, a wide range of PCR techniques have been described for IK, including species-specific PCR,^21^ semi-nested/nested PCR,^30^ touchdown PCR,^31^ multiplex PCR,^32^ real-time/quantitative PCR (rt-/q-PCR),^33,34^ and broad-range 16S/18S PCR.^14,18,19,21,35,36^ To the best of our knowledge, this is the first study that had directly compared the performance of three different microbiological investigations, namely direct culture, indirect culture, and 16S/18S rRNA PCR, for diagnosing IK. We observed comparable diagnostic yield among all three tests (41.9-53.0%), with moderate-to-substantial inter-test agreement between culture and PCR (especially in more severe infection cases).

So far, there were limited studies in the UK^14,21^ and few outside the UK^19,26,35,37^ that had evaluated the performance of culture and 16S rRNA PCR for bacterial keratitis. When compared to Somerville et al. study,^14^ we observed a considerably higher diagnostic yield for both culture (53.0% vs. 23.0%) and PCR (41.9% vs. 26.0%) methods in our study. The discrepancy might be attributed to different proportion of patients with prior antimicrobial use, heterogeneous patient cohorts (with different presenting severity of infection), and corneal sampling techniques. There was a significantly higher use of prior antimicrobial reported in Somerville et al. study (34.0%) as opposed to our study (8.1%), which might have negatively impacted on the diagnostic yield in their study. Similar to their study, our study did not demonstrate any superiority in the diagnostic performance of PCR over culture in cases with prior antimicrobial use. Although the performance of PCR is reported to be less affected by prior use of antimicrobials than microbiological culture, the impact may still be clinically significant, particularly when there is a long interval between antimicrobial exposure and sample collection.^38,39^ In addition, there is a significantly lower infectious biomass in IK (i.e. the entire corneal surface area is only 1.3 cm^2^) as opposed to systemic infections (e.g. pneumonia or sepsis), and the constant tear drainage and increased tearing in IK setting might further dilute the microbial load, negatively impacting on the diagnostic yield.^3^

Since the publication of our previous studies,^20,23^ we had also implemented further training on corneal sampling and a change in the sampling technique/instrument (from needle/blade to flocked swabs, known to improve the diagnostic yield),^24,40^ which might have accounted for the improvement in the culture yield by 15% within our practice (from 37.7% to 53.0%). In addition, previous studies, including our Nottingham studies, have shown that culture positivity is significantly influenced by the initial severity of the infection.^4,5,41^ As the clinical severity of IK was not reported in Somerville et al. study, direct comparison with their study was not possible from this aspect.

Based on the reference standard used (either direct culture standard or composite reference standard), 16S rRNA PCR was shown to exhibit a good sensitivity (73.5-74.3%) and specificity (90.3-100%) for diagnosing bacterial keratitis in our study. This was comparable to the previous findings reported in the literature, with a sensitivity of 63.6-100% and specificity of 67.5-100%.^3^ Interestingly, a recent UK study by Hoffman et al. reported a significantly lower sensitivity in PCR (25.0%) than microbiological culture (95.6%) for diagnosing bacterial keratitis.^21^ As both studies utilized the same commercial company (MicroPathology Ltd, Coventry, UK) for PCR-Sanger sequencing, the higher sensitivity of PCR observed in our study is likely attributed to the differences in corneal sampling techniques, sample transportation method, and inclusion criteria / clinical threshold for performing corneal sampling for PCR.

One of the inherent issues with high sensitivity of PCR is that the test can produce false positive results as the test picks up environmental or internal contaminants (e.g. from latent host DNA);^19,37,42^ for instance, some universal primers of 16S rRNA genes may amplify eukaryotic rRNA genes non-specifically, producing false positive results.^43^ Somerville et al.^14^ demonstrated 16S rRNA amplicons in negative controls, highlighting the possibility of background contamination resulting in false positives. To address this issue, the PCR total cycle number was set at 30 (but not higher) to avoid false positive results, which could potentially explain the significantly lower rate of *Propionibacterium spp.* (a common ocular surface commensal) diagnosed by the PCR (2.8%) when compared to the direct and indirect culture methods (12.2-22.2%). Despite having the lowest but comparable sensitivity, 16S rRNA PCR was able to improve the diagnostic yield in direct culture-negative cases by ∼10%, highlighting its adjuvant role for the diagnosis of IK.

On the other hand, some studies have shown that PCR has a higher yield than culture/smear, which might have been due to the differences in types of infection (e.g. ocular vs. non-ocular infections), sampling techniques and sample transport/storage methods, laboratory techniques (e.g. choice of universal primers, extraction and amplification techniques), and patient cohort (e.g. varied clinical severity and types of infectious keratitis).^16,19,26,37,39^ Although our study was set out to include fungal keratitis cases, none of the culture or PCR yielded any fungal pathogen, limiting the interpretation of 18S rRNA PCR for diagnosing fungal keratitis. That said, the lack of false positive results and the perfect concordance between culture and 18S rRNA PCR suggests a high specificity of this test for ruling out fungal keratitis (though the sensitivity remains to be elucidated in our patient cohort).

In addition, we observed a very high percent agreement (>80%) between culture and PCR. Although good PCR-culture concordance (77.0-90.6%) has been previously reported,^14,35^ low concordance has also been noted in some retrospective studies.^21,44^ It is noteworthy to highlight that almost all studies reported test concordance in the form of percent agreement. While percent agreement serves as an easy way to interpret inter-test agreement rate, such analysis does not consider chance agreement and may erroneously overestimate inter-test agreement.^29,45^ In view of this issue, we performed additional Cohen’s *k* analysis, which showed substantial pairwise agreement on microbiological results among all three tests (mean k=0.67-0.72). To dissect the potential reasons influencing test concordance, we further performed clinico-microbiological correlation, which showed that positive culture-PCR concordance was most likely to be achieved in more severe IK cases, corroborating the findings observed in the Shimizu et al. study.^39^ This suggests that 16S rRNA PCR may have more additional clinical values in culture-negative IK cases or those that have lesser severity of infection where it can provide additional information that may not be detected by the culture method.

Another strength of this comparative study lies in the inclusion of an indirect culture group. The reasons for including an indirect culture group were twofold. First, we aimed to explore the diagnostic potential of indirect culture in replacing the direct culture method (which was the gold standard in our practice and many others) as indirect culture is more time-efficient since it only requires one sampling at the front-line ophthalmic setting, with further sub-inoculation being done at the microbiology laboratory. Our study demonstrated similar diagnostic performance and inter-test concordance between direct and indirect culture, supporting its potential use in a busy clinical setting. This also supports the findings of previous similar studies.^8,9^ Secondly, both indirect culture and PCR test were performed based on the same sample from the inoculated transport medium, allowing a direct comparison without any influence of the sampling sequence. Although a few studies have demonstrated that the sequence of sampling did not affect the diagnostic yield,^14^ the theoretical impact and concern of having less infectious biomass in later samplings still exist. Therefore, having an indirect culture group (which utilized the same inoculated sample for PCR) addressed this issue.

One of the limitations of this study was the moderate sample size, an issue that was commonly noted in many previous studies.^14,19,26,37^ However, the number of cases included in our study (n=65 samples per test group) satisfied the minimum required sample size (n=61 per test group) based on our calculation. Our study observed a moderate diagnostic yield in both culture and PCR methods (ranged 40-50%), which might be related to the inclusion of less severe infection cases (∼60% of the cases have an infiltrate size of <3mm). Nonetheless, the diagnostic yield was higher than our previous study (37.7%),^20^ suggesting an improvement of our sampling technique. A cost-benefit analysis of PCR use in IK diagnosis would also be beneficial as earlier accurate diagnosis could improve clinical outcomes and reduce the risk of complication and need for surgical interventions. For example, Knight et al.^46^ investigated three different PCR and culture combinations to diagnose *Enterobacteriaceae* infection and found the combination of PCR and culture to be the most cost-effective. In the future, multiplex PCR or metagenomic next generation sequencing may also further improve the diagnosis of IK, though its routine use in clinic is currently limited by the cost and the availability of technical expertise and resources.^47–51^

In conclusion, this study demonstrated good diagnostic performance of all three different microbiological diagnostic modalities, namely direct culture, indirect culture and PCR. We also highlight the potential adjuvant role of 16S rRNA PCR for bacterial keratitis, particularly in culture-negative and less severe IK cases. However, larger studies and cost-benefit analysis are required to determine its role for routine use in future clinical practice.

## Data Availability

The authors confirm that the data supporting the findings of this study are available within the article (and its supplementary materials).

