## Supplementary Table 1 for "Microbiological Culture Versus 16S/18S Ribosomal RNA PCR-Sanger Sequencing for Infectious Keratitis: A Three-Arm, Diagnostic Cross-Sectional Study"

Supplementary Table 1. A 2x2 cross-tabulation for calculating sensitivity and specificity.

|  | | **Disease status (based on reference standard)** | |
| --- | --- | --- | --- |
|  |  | Yes | No |
| **Index test** | Positive | True positive (TP) | False positive (FP) |
|  | Negative | False negative (FN) | True negative (TN) |
| **Sensitivity and specificity** | | Sensitivity:  $=\frac{TP}{TP+FN}$ | Specificity:  $=\frac{TN}{FP+TN}$ |
