## Supplementary material for "Microbiological Culture Versus 16S/18S Ribosomal RNA PCR-Sanger Sequencing for Infectious Keratitis: A Three-Arm, Diagnostic Cross-Sectional Study": Table 1

Table 1. Patient and clinical characteristics of suspected bacterial or fungal keratitis at Queen’s Medical Centre, Nottingham, UK, between June 2021 and September 2022.

| Parameters | **All cases (N=81)**  N (%) | **Culture/PCR positive (N=47)**  N (%) | **Culture/PCR negative (N=34)**  N (%) | **P-value*** |
| --- | --- | --- | --- | --- |
| Age (years)  Female gender  Right eye  Duration of symptoms  Risk factors**  Contact lens wear  Ocular surface disease  Topical steroids  Trauma  Prior corneal surgery  Systemic immunosuppression***  Presenting CDVA (logMAR)  Epithelial defect size^#^  Small (<3.0mm)  Moderate (3.1-6.0mm)  Large (>6.0mm)  Infiltrate size^#^  Small (<3.0mm)  Moderate (3.1-6.0mm)  Large (>6.0mm)  Location^#^  Peripheral  Paracentral  Central  Presence of hypopyon | 49.8 ± 22.7  41 (50.6)  41 (50.6)  4.8 ± 8.0  43 (53.1)  34 (42.0)  14 (17.3)  10 (12.3)  10 (12.3)  9 (11.1)  0.93 ± 0.93  48 (59.3)  18 (22.2)  9 (11.1)  51 (63.0)  20 (24.7)  5 (6.2)  13 (16.0)  37 (45.7)  28 (34.6)  16 (19.8) | 50.3 ± 23.0  24 (51.1)  26 (55.3)  3.9 ± 5.0  28 (59.6)  21 (44.7)  11 (23.4)  3 (6.4)  6 (12.8)  6 (12.8)  0.97 ± 0.93  24 (51.1)  13 (27.7)  7 (14.9)  30 (63.8)  10 (21.3)  5 (10.6)  5 (10.6)  22 (46.9)  18 (38.3)  13 (27.7) | 48.8 ± 22.6  17 (50.0)  15 (44.1)  6.2 ± 10.9  15 (44.1)  13 (38.2)  3 (8.8)  7 (20.6)  4 (11.8)  3 (8.8)  0.84 ± 0.91  24 (70.6)  5 (14.7)  2 (5.9)  21 (61.8)  10 (29.4)  0 (0.0)  8 (23.5)  15 (18.5)  10 (29.4)  3 (8.8) | 0.77  0.92  0.32  0.21  0.28  0.55  0.12  0.13  0.28  0.036 |

PCR = 16S/18S ribosomal ribonucleic acid (rRNA) polymerase chain reaction; CDVA = Corrected-distance-visual acuity

*Statistical comparison was made between culture/PCR-positive (patients were positive on at least one test) and culture/PCR-negative (patients were negative on all tests) cases.

**Some patients have more than one risk factor.

***Includes use of systemic immunosuppressive drugs, diabetes, and immunodeficiency.

^#^Some missing data on the corneal ulcer characteristics.

Continuous variables are presented as mean ± standard deviation (SD).
