## Supplementary material for "Microbiological Culture Versus 16S/18S Ribosomal RNA PCR-Sanger Sequencing for Infectious Keratitis: A Three-Arm, Diagnostic Cross-Sectional Study": Table 2

Table 2. A summary of the causative organisms isolated in patients presenting with presumed infectious keratitis to the Queen’s Medical Centre, Nottingham, UK.

| **Organisms*** | **Total****  **(N=66)**  N% | **Direct Culture (N=41)**  N (%) | **Indirect Culture (N=54)**  N (%) | **PCR**  **(N=36)**  N (%) | **P-value^***^** |
| --- | --- | --- | --- | --- | --- |
| Gram-positive | 38 (57.6) | 19 (46.3) | 32 (59.3) | 14 (38.9) | 0.15 |
| CoNS | 15 (22.7) | 5 (12.2) | 12 (22.2) | 7 (19.4) | 0.45 |
| *Propionibacterium* *spp.* | 13 (19.7) | 5 (12.2) | 12 (22.2) | 1 (2.8) | 0.030 |
| *Staphylococcus aureus* | 3 (4.6) | 5 (12.2) | 3 (5.6) | 2 (5.6) | 0.41 |
| *Streptococci spp*. | 3 (4.6) | 3 (7.3) | 3 (5.6) | 2 (5.6) | 0.93 |
| Others^#^ | 4 (6.1) | 1 (2.4) | 2 (3.7) | 2 (5.6) | 0.77 |
| Gram-negative | 28 (42.4) | 22 (53.7) | 22 (40.7) | 22 (61.1) | 0.15 |
| *Pseudomonas aeruginosa* | 14 (21.2) | 13 (21.7) | 11 (20.4) | 10 (27.8) | 0.44 |
| *Serratia spp.* | 6 (9.1) | 3 (7.3) | 6 (11.1) | 6 (16.7) | 0.44 |
| *Moraxella spp.* | 4 (6.1) | 3 (7.3) | 3 (5.6) | 3 (8.3) | 0.87 |
| Others^$^ | 4 (6.1) | 3 (7.3) | 2 (3.7) | 3 (8.3) | 0.62 |

PCR = 16S/18S rRNA polymerase chain reaction; CoNS = Coagulase-negative Staphylococci

* The organisms presented here are based on 66 cases of direct culture, 85 indirect culture, and 86 cases of PCR. The “N” refers to the number of organisms in each test group. The numbers are higher that the diagnostic yield of each respective test as some cases are caused by more than 1 organism.

** The total was based on the causative organisms isolated by all three tests. For example, if a case was positive for two different organisms on either the same or different tests, both organisms were included for that case. However, the same organism isolated by different methods in each case was considered as one organism.

*** Comparison of the causative organisms was made among the three test groups, based on the total number of organisms in each group.

^#^ These included *Abiotrophia defectiva*, *Bacillus spp.,* and *Corynebacterium spp*.

^$^ These included *Citrobacter* *koseri*, *Acinetobacter spp.,* and *Hemophilus influenzae.*
