## Supplementary material for "Microbiological Culture Versus 16S/18S Ribosomal RNA PCR-Sanger Sequencing for Infectious Keratitis: A Three-Arm, Diagnostic Cross-Sectional Study": Table 3

Table 3. Diagnostic performance of indirect culture and 16S/18S rRNA PCR against direct culture (reference standard).

|  | | **Direct culture** | | |  |  | | **Direct culture** | | |
| --- | --- | --- | --- | --- | --- | --- | --- | --- | --- | --- |
|  |  | P | N | Total |  |  |  | P | N | Total |
| **Indirect culture** | P | 29 | 4 | 33 |  | **PCR** | P | 26 | 3 | 29 |
|  | N | 5 | 27 | 32 |  |  | N | 9 | 28 | 37 |
|  | Total | 34 | 31 | 65 |  |  | Total | 35 | 31 | 66 |
| **Performance** | | Sn = 85.3% | Sp = 87.1% |  |  | Performance | | Sn = 74.3% | Sp = 90.3% |  |

P = Positive microbiological result; N = Negative microbiological result; PCR = 16S/18S rRNA polymerase chain reaction; Sn = Sensitivity; Sp = Specificity
