## Supplementary material for "Microbiological Culture Versus 16S/18S Ribosomal RNA PCR-Sanger Sequencing for Infectious Keratitis: A Three-Arm, Diagnostic Cross-Sectional Study": Table 4

Table 4. Diagnostic performance of direct culture, indirect culture, and 16S/18S rRNA PCR, based on the composite reference standard (CRS).

|  | | **CRS** | | | |  | |  | | **CRS** | | | |  |  | | **CRS** | | | |
| --- | --- | --- | --- | --- | --- | --- | --- | --- | --- | --- | --- | --- | --- | --- | --- | --- | --- | --- | --- | --- |
|  |  | P | N | Total |  | |  | | | P | N | Total |  | |  | | | P | N | Total |
| **DC** | P | 35 | 0 | 35 |  | | **IC** | | P | 41 | 0 | 41 |  | | **PCR** | P | | 36 | 0 | 36 |
|  | N | 5 | 26 | 31 |  | |  |  | N | 7 | 37 | 44 |  | |  | N | | 13 | 37 | 50 |
|  | Total | 40 | 26 | 66 |  | |  |  | Total | 48 | 37 | 85 |  | |  | Total | | 49 | 37 | 86 |
| **Performance** | | Sn = 85.3% | Sp = 100% |  |  | | **Performance** | | | Sn = 85.4% | Sp = 100% |  |  | | **Performance** | | | Sn = 73.5% | Sp = 100% |  |

DC = Direct culture; IC = Indirect culture; PCR = 16S/18S rRNA polymerase chain reaction; P = Positive microbiological result; N = Negative microbiological result; Sn = Sensitivity; Sp = Specificity
