## Supplementary material for "Microbiological Culture Versus 16S/18S Ribosomal RNA PCR-Sanger Sequencing for Infectious Keratitis: A Three-Arm, Diagnostic Cross-Sectional Study": Table 5

Table 5. Concordance between microbiological investigations for diagnosing infectious keratitis (IK).

| **Comparison** | **Number of cases** | **Percent agreement,**  N (%) | **Cohen’s Kappa (*k*),**  Mean ± SE (95% CI) |
| --- | --- | --- | --- |
| *Concordance at microbiological detection level** | | | |
| DC versus IC | 65 | 56 (86.2%) | 0.72 ± 0.09 (0.55-0.89) |
| DC versus PCR | 66 | 54 (81.8%) | 0.67 ± 0.09 (0.49-0.84) |
| IC versus PCR | 85 | 73 (85.9%) | 0.69 ± 0.08 (0.54-0.84) |
| *Concordance at organism genus level (in positive cases only)*** | | | |
| DC versus IC | 65 | 55 (84.6%) | 0.69 ± 0.09 (0.52-0.87) |
| DC versus PCR | 66 | 51 (77.3%) | 0.55 ± 0.10 (0.36-0.74) |
| IC versus PCR | 85 | 70 (82.4%) | 0.62 ± 0.08 (0.46-0.79) |

DC = Direct culture; IC = Indirect culture; PCR = 16S/18S rRNA polymerase chain reaction; CI = Confidence interval

*Concordance is considered based on the microbiological detection result (i.e. positive or negative test result). Concordance is achieved if detection was positive on positive on both tests, regardless of the causative organism(s).

**Concordance is considered based on the organism’s genus result. Concordance is achieved if the same organism (at least one) is isolated by both comparing techniques.
