## Supplementary material for "Microbiological Culture Versus 16S/18S Ribosomal RNA PCR-Sanger Sequencing for Infectious Keratitis: A Three-Arm, Diagnostic Cross-Sectional Study": Table 6

Table 6. Examination of potential factors influencing the concordance between direct culture and 16S/18S rRNA PCR results in culture- or PCR-positive infectious keratitis (IK).

| **Parameters** | **All cases (N=38)**  N (%) | **Culture-PCR matched** **(N=26)**  N (%) | **Culture-PCR unmatched (N=12)**  N (%) | **P-value*** |
| --- | --- | --- | --- | --- |
| Age (years) | 52.3 ± 21.5 | 53.3 ± 20.4 | 50.2 ± 24.7 | 0.71 |
| Female gender | 20 (52.6) | 14 (53.8) | 6 (50.0) | 1.0 |
| Right eye | 20 (52.6) | 15 (57.7) | 5 (41.7) | 0.49 |
| Duration of symptoms, days | 5 ± 9.1 | 5 ± 11.0 | 3 ± 2.6 | 0.41 |
| Presenting CDVA, logMAR | 1.08 ± 1.03 | 1.29 ± 1.06 | 0.62 ± 0.81 | 0.040 |
| Epithelial defect size, mm^$^  Small (≤3.0)  Moderate/large (>3.0) | 16 (44.4)  20 (55.6) | 9 (36.0)  16 (74.0) | 7 (63.6)  4 (36.4) | 0.16 |
| Infiltrate size, mm^$^  Small (≤3.0)  Moderate/large (>3.0) | 24 (63.2)  13 (36.8) | 15 (60.0)  10 (40.0) | 9 (75.0)  3 (25.0) | 0.48 |
| Location^$^  Peripheral/paracentral  Central | 21 (58.3)  15 (41.7) | 13 (52.0)  12 (48.0) | 8 (72.7)  3 (27.3) | 0.30 |
| Presence of hypopyon | 13 (34.2) | 11 (42.3) | 2 (16.7) | 0.16 |

*Comparison was made between culture-PCR matched and unmatched groups, using unpaired T-test (for continuous variables) and Chi-square or Fisher exact test (for categorical variables where appropriate).

^$^A few missing data on the corneal ulcer characteristics.
